# RAB5A expression is a predictive biomarker for trastuzumab emtansine in breast cancer

**DOI:** 10.1101/2021.07.06.21255069

**Authors:** Olav Engebraaten, Christina Yau, Kristian Berg, Elin Borgen, Øystein Garred, Maria E.B. Berstad, Ane S.V. Fremstedal, Angela De Michele, Laura van’t Veer, Laura Esserman, Anette Weyergang

**Affiliations:** Department of Oncology, Oslo University Hospital, Oslo, Norway and Institute of Clinical Medicine, University of Oslo, Norway; Institute for Cancer Research, Norwegian Radium Hospital, Oslo University Hospital, Norway; Department of Surgery, University of California San Francisco, California, United States; Department of Pharmacy, University of Oslo, Norway; Department of Pathology, Oslo University Hospital, Oslo Norway; Department of Medicine, Perelman school of Medicine, University of Pennsylvania, United States

## Abstract

**PURPOSE:** Targeted therapeutics strongly depends on validated biomarkers in order to select patients most likely to benefit from the treatment. HER2 serves as a predictive biomarker for HER2-targeted tyrosine kinase inhibitors and monoclonal antibodies. HER2 may, however, also be utilized as a transport gate for delivery of cytotoxic agents into the cell, such as for HER2-targeted antibody drug conjugates (ADCs; e.g. trastuzumab emtansine (T-DM1)). The predictive biomarkers for such ADCs may be more complex, also reflecting the intracellular transport.

**METHODS:** Five HER2-positive breast and ovarian cancer cell lines were evaluated with respect to T-DM1 sensitivity and correlated to the expression levels of proteins involved in endocytic trafficking including RAB4A, RAB5A and RAB11A, with possible impact on ADC pharmacology. The results were confirmed in a clinical cohort consisting of patients from the adaptive breast cancer clinical trial I-SPY2 where pathological complete response (pCR) was correlated to the RNA expression level of RAB4A, RAB5A and RAB11A. A subset of the clinical KAMILLA trial including 19 patients was used as a verification cohort where semi-quantitative IHC of RAB5A was correlated to progression free survival (PFS).

**RESULTS:** The early endosome marker RAB5A, was found to correlate positively to T-DM1 sensitivity in the cell line panel. Correlation between RAB5A expression and T-DM1 sensitivity (pCR) was confirmed in patients treated with trastuzumab emtansine/pertuzumab in the I-SPY2 trial, but not in the trastuzumab/paclitaxel control arm. The clinical correlation was verified in the patients from the KAMILLA trial where semi-quantitative RAB5A IHC staining correlated significantly positive to PFS.

**CONCLUSION:** The present results indicate that RAB5A is a predictive biomarker for T-DM1 and outline, for the first time, proteins involved in endocytic trafficking as predictive biomarkers for ADCs.

## INTRODUCTION

The increased focus on personalized medicine has together with our increasing knowledge in cancer biology revealed a great potential for the use of biomarkers in cancer treatment. A number of different biomarkers are already incorporated in clinical practice to predict patient survival, select an appropriate therapy or monitor disease progression^1^. Predictive biomarkers enable careful selection of those patients most likely to benefit from a specific treatment, and hence, such knowledge is crucial in order to rationally exploit current and future high-cost targeted cancer therapeutics. HER2 (ERBB2) is a validated biomarker in breast cancer and HER2 gene amplification or protein over-expression is found in ∼20 % of newly diagnosed breast cancer patients^2,3^. HER2 is utilized as a predictive biomarker for treatment with HER2-targeting monoclonal antibodies (mAbs) (trastuzumab and pertuzumab) and tyrosine kinase inhibitors (TKIs) (lapatinib and afatinib)^3^. The pharmacological effects of HER2-targeted mAbs and TKIs are a direct consequence of drug-target interaction and include antibody-dependent cellular cytotoxicity (ADCC) (mAbs), HER2 down regulation and inhibition of growth promoting signaling^4-6^. The ability of HER2 to undergo receptor-mediated endocytosis also makes this transmembrane protein a candidate for delivery of cytotoxic agents into the cancer cells. This has indeed been exemplified by the antibody-drug conjugate (ADC) trastuzumab emtansine (T-DM1)^7,8^ which received FDA approval for treatment of metastatic breast cancer in 2013.

T-DM1 consists of trastuzumab linked by a thioether (N-maleimidomethyl cyclohexane-1-carboxylate (MCC)) to the highly cytotoxic maytansine-derived drug, DM-1^9^. Upon administration, T-DM1 binds to HER2 and is taken into the cell by HER2-mediated endocytosis. Proteolytic degradation of the trastuzumab-component within the endo/lysosomal pathway is postulated as the mechanism for cytosolic release of DM1 which subsequently induces microtubule destabilization and cell death^10,11^. T-DM1 therefore induces a cytotoxic mechanism of action within the cell in addition to the pharmacological effects generated by its trastuzumab-component.

The action mechanisms of T-DM1 are clearly more complex than that of HER2-targeting mAbs^12,13^ and TKIs and we have evaluated if this is reflected in the biomarkers that can be used to predict drug-response. Candidate biomarkers for T-DM1 efficacy have, until now, focused on HER2 and its downstream signaling in addition to HER3^14,15^, and little is known about the impact of proteins involved in endocytosis, endocytic vesicle transport and exocytosis. The present study is the first report addressing proteins involved in intracellular trafficking as predictive biomarkers for T-DM1 treatment response.

## 2. MATERIALS AND METHODS

### 2.1 Cells and culturing

Five HER2-expressing human cell lines were used in this study; the breast cancer cell lines SK-BR-3 (HTB-30), AU-565 (CRL-2351), HCC1954 (CRL-2338) and MDA-MB-453 (HTB-131) and the ovarian cancer cell line SKOV-3 (HTB-77). The human breast cancer cell line MDA-MB-231 was used as a negative control for HER2 expression. All cell lines were obtained from American Type Culture Collection (Manassas, VA, USA), except SK-BR-3, kindly provided by the Department of Biochemistry at Institute for Cancer Research, Norwegian Radium Hospital. All cell lines were used between passage number 3 and 25 to avoid changes in the cell line characteristics with time, and the cells were routinely checked for Mycoplasma infections. SK-BR-3 and SKOV-3 cells were cultured in McCoy’s 5A medium, AU-565, HCC1954 and MDA-MB-231 cells in RPMI-1640 medium (both obtained from Sigma-Aldrich, St. Louis, MO, USA), while MDA-MB-453 were cultured in Leibovitz’s L-15 medium (Lonza, Verviers, Belgium). All media were supplemented as previously described ^16^.

### 2.2 Cytotoxicity experiments

Cells were seeded at 8×10^3^ (SK-BR-3), 1.8×10^3^ (SKOV-3), 6×10^3^ (AU-565), 4×10^3^(HCC1954) or 1×10^4^ cells/well (MDA-MB-453) in 96-well plates (Nunc, Roskilde, Denmark) and allowed to attach overnight. The cells were then incubated with trastuzumab (Herceptin®, Roche, Basel, Switzerland) or T-DM1 (ado-trastuzumab emtansine, Kadcyla®, Genentech, San Francisco, CA, USA) at increasing concentrations for 72 hrs, after which cell viability was assessed by the MTT assay as previously described^17^. IC_50_ values were calculated from sigmoidal curves (fit model: a/(1 + exp(-(*x* - *x*0) / *b*)) generated in SigmaPlot 14 with integrated SigmaStat (Synstat Software, Inc, Jan Jose, Ca, USA).

### 2.3 Western blot analysis

Total cell extracts were obtained from ∼80% confluent cells seeded in 6 well plates. The cells were washed once with PBS and collected in 700μl PBS with a cell scrape on ice before they were subjected to centrifugation at 1000xg at 4°C for 5 min. The supernatant was removed and the pellet kept at -80 °C until lysis. The cell pellets were lysed with 50-150μl lysis buffer (150mM NaCl, 50mM Tris pH 7.5, 0.1% SDS) including Halt protease and phosphatase inhibitor cocktail (Thermo Fisher Scientific) on ice for 15-30 min. The lysates were then sonicated and spun down at 12.000xg at 4°C for 15 min. The protein concentration in the supernatants was assessed by the Bio-Rad Protein Assay Dye Reagent Concentrate (Bio-Rad Laboratories, Ca USA) before the lysate was transferred to new tubes and stored at -80 °C until SDS-PAGE and Western blotting as previously described^18^. Cellular protein expression was detected using HER2 (#2165) antibody from Cell Signaling Technology (Danvers, MA, USA), RAB5A (PA5-29022), RAB11A (71-5300) and RAB4A (PA3-912) antibody from Thermo Fisher Scientific (Rockford, IL, USA). Protein expression was correlated to γ-tubulin as detected by an antibody (#T6557) from Sigma-Aldrich. HRP linked α-rabbit (#7074) and α-mouse (#7076) antibodies from Cell Signaling Technology were used as secondary antibodies. Supersignal West Dura Extended duration Substrate (Thermo Scientific, Rockford, IL, USA) and ChemiDoc^™^ densitometer (Bio-Rad) was used for the detection of protein bands on the membrane. ImageLab 4.1 (Bio-Rad) (software) was used for quantification of protein expression. The expression of each protein was calculated relative to the highest expressing cell line.

### 2.4 *In vitro* correlation analysis

The relative expression of HER2 RAB4A, RAB5A and RAB11A in the cell lines was plotted against the cell line sensitivity towards T-DM1, as measured by 1/IC_50_ (concentration inhibiting the viability by 50%) and a linear regression including the R^2^ value was assessed using SigmaPlot 15 with integrated SigmaStat.

### 2.5 I-SPY 2 TRIAL

The I-SPY 2 TRIAL is a multicenter, open-label adaptive neoadjuvant platform trial for women with breast cancer (> 2,5 cm clinically and > 2 cm by imaging) including biomarker assessments (based on HER2 status, estrogen and progesterone receptors, and a 70-gene assay (MammaPrint, Agendia)) prior to inclusion. Core biopsy samples are secured for RNA expression analyses, and one of the study aims of I-SPY 2 is to test and validate biomarkers for new drugs^19^ (NCT01042379). T-DM1 + pertuzumab (T-DM1+P) were one of the novel combinations evaluated for efficacy in I-SPY 2, against a trastuzumab + paclitaxel (TH) control (manuscript submitted). In the present study RAB5A, RAB4A and RAB11A expression was evaluated as specific predictors of pathologic complete response (pCR) to T-DM1 + pertuzumab. Although trastuzumab + paclitaxel + pertuzumab was also evaluated as an experimental regimen over the same period, formal comparisons between two experimental arms are contractually prohibited. Therefore, a separate qualifying biomarker analysis was performed on patients receiving trastuzumab + paclitaxel + pertuzumab to evaluate whether these biomarkers associated with response to pertuzumab.

#### Expression data

All I-SPY 2 samples were analyzed on one of two Agilent custom arrays (the 15746 and 32627 designs). All samples in the T-DM1+pertuzumab and trastuzumab + paclitaxel + pertuzumab arm was assayed on the 32627 arrays, while the trastuzumab+paclitaxel arm was split between the platforms, with 22 samples on the older 15746 platform and 9 samples on the 32627 array. To combine data across the two designs, the probe annotation of the 15746 platform was updated (September 2016); and for each platform, collapsed normalized expression data by averaging such that genes represented by multiple probes are computed as the average across probes. The ComBat algorithm was then applied to adjust for platform-biases and combine the data from the two platforms. This procedure was performed for the pre-treatment data of the first 880 I-SPY 2 patients irrespective of experimental arm. Normalized, platform-corrected pre-treatment expression levels of RAB5A, RAB4A and RAB11A from patients in the T-DM1+pertuzumab arm (n=52), the trastuzumab + paclitaxel control (n=31) and the trastuzumab + paclitaxel + pertuzumab (n=44) (Supplemental Table 1) were evaluated individually for association with pCR.

#### Qualifying biomarker analysis

All I-SPY qualifying biomarker analyses follow similar multi-step pre-specified analysis plans^20-22^. Associations with pCR were first assessed within each arm with logistic modelling and significance assessment using the likelihood ratio test. As well, the interaction between biomarker and treatment was evaluated using a logistic model fitted to data from the T-DM1+pertuzumab and control arms. These analyses were also performed adjusting for HR status as a covariate.

In a second step, an optimal dichotomizing threshold was determined for biomarkers which specifically associate with response in the T-DM1+pertuzumab but not the control arm and have a significant (p<0.05) biomarker x treatment interaction, using a Monte-Carlo 2-fold cross-validation procedure. Specifically, for 100 iterations, half of the cases were randomly selected, balancing for treatment arm and pCR status, as training set. Every value between the 10th and 90th percentile were considered as a potential threshold to dichotomize the training set into “High” vs. “Low” RAB5A expressing groups; and fit a series of logistic regression models to assess the biomarker x treatment interaction. The threshold which minimizes the likelihood-ratio (LR) test p value for the interaction term in the training set was selected and used to dichotomize the test set, and assess the significance of the biomarker x treatment interaction in the test set. The LR p values across the 100 test sets were then combined using the logit method; and the threshold yielding the minimum combined LR test p value was selected. In the final step, the optimal threshold identified to dichotomize patients into RAB5A High and RAB5A Low groups was used to calculated the Bayesian estimated PCR probability in the T-DM1+pertuzumab and control arms using a Bayesian co-variate adjusted logistic model similar to the standard one I-SPY 2 uses to evaluate agent efficacy (but without time-adjustment).

#### Establishment of Receiver operating characteristic (ROC) curves

The patients in each treatment group were sorted according to RAB5A expression. ROC curves were established to visualize how the expression level correlated with pCR. The SigmaPlot software with integrated SigmaStat was used for the establishment and analysis of ROC curves. The T-DM1+P curve was also used to identify a threshold for RAB5A expression to dichotomize patients into RAB5A -high and -low.

### 2.6 KAMILLA TRIAL

The KAMILLA trial (NCT01702571) is an international multicenter single-arm, open-label phase IIIb safety study of T-DM1 including patients with HER2-positive advanced breast cancer with progression after prior treatment with chemotherapy and a HER2 directed agent for metastatic disease^23^. The Norwegian Radium Hospital, OUS was included as one of the centers recruiting patients in the Kamilla study, and primary FFPE biopsies were available for 19 of the patients. The patients received T-DM1 until unacceptable toxicity, withdrawal or disease progression. The data from the patients included in the current study were retrieved directly from the medical records of the patients. Three patients, all with a progression free survival of more than 50 weeks, either stopped treatment due to toxicity or were lost to follow-up. All other patients discontinued treatment due to progression of the disease. In the present study these primary biopsies were subjected to IHC using an anti-RAB5A from Abcam (ab 109534; rabbit IgG, clone EPR5438, diluted 1:1600; incubation 30 minutes on a Dako Autostainer platform. Antigen retrieval was performed using Dako’s High pH solution in a PT link, and as detection system was used Dako’s EnVision Flex+ system with rabbit linker (K8009). Slides from FFPE blocks from SKBR3 and Skov3 cell line cells were included in the staining runs as positive and negative controls, respectively. The RAB5A staining, localized to the cytoplasm and/or membrane of the tumor cells, was quantified using the Allred Scoring system, where the staining is evaluated based on both color intensity (0— Negative, 1—Weak, 2—Intermediate, 3—Strong) and proportion of stained cells (0—No positive, 1—≤ 1% positive, 2—1–10% positive, 3—11–33% positive, 4—34–66% positive, 5—67–100% positive)^24,25^. RAB5A expression, as measured by Allred score was correlated to progression-free survival (PFS) data.

### 2.7 Ethics oversight and consent

Both the I-SPY 2 and Kamilla study complied with all relevant ethical regulations for work with human participants. The I-SPY2 study was approved by IRB boards at the participating study sites:

^*^Institution: University of California San Diego Moores Cancer Center

Name and address IRB: University of California, San Diego Human Research

Protections Program Institutional Review Boards (Attn: Human Research Protections

Program (HRPP) Altman Clinical and Translational Institute, Level 2 9452 Medical Center Drive La Jolla, CA 92093)

^*^Institution: Georgetown University Lombardi Cancer Center

Name and address: MedStar Health Research Institute-Georgetown University Oncology

Institutional Review Board (Medical-Dental Building, SW104, 3900 Reservoir Road NW, Washington, DC 20057)

^*^Institution: Loyola University Chicago Stritch School of Medicine, Cardinal Bernardin Cancer Center

Name and address: Loyola University Chicago Health Sciences Division Institutional Review Board for the Protection of Human Subjects (2160 South First Avenue Maywood, IL 60153)

^*^Institution: University of California, San Francisco, Helen Diller Family of Comprehensive Cancer Center

Name and address: UCSF Human Research Protection Program Institutional Review Board (490 Illinois Street, Floor 6, San Francisco, CA 94143)

^*^Institution: University of Texas, Southwestern Medical Center Simmons Comprehensive Cancer Center

Name and address: UT Southwestern IRB (5323 Harry Hines Blvd. Dallas, TX 75390)

^*^Institution: H. Lee Moffitt Cancer Center and Research Institute

Name and address: Chesapeake IRB (3181 SW Sam Jackson Park Road - L106RI Portland, OR 97239-3098)

^*^Institution: Oregon Health and Science University Knight Cancer Institute

Name and address: Oregon Health & Science University Research Integrity Office IRB (3181 SW Sam Jackson Park Road - L106RI, Portland, OR 97239-3098

^*^Institution: Mayo Clinic Breast Cancer Center - Rochester

Name and address: Mayo Clinic Institutional Review Boards (201 Building, Room 4-60, 200 First St. SW, Rochester, MN 55905)

^*^Institution: University of Pennsylvania, Abramson Cancer Center

Name and address: University of Pennsylvania Office of Regulatory Affairs Institutional Review Board (3624 Market St., Suite 301 S, Philadelphia, PA 19104-6006)

^*^Institution: University of Alabama at Birmingham Comprehensive Cancer Center

Name and address: The University of Alabama at Birmingham Office of the Institutional Review Board for Human Use (470 Administration Building, 701 20th Street South, Birmingham, AL 35294-0104)

^*^Institution: University of Minnesota, Masonic Cancer Center

Name and address: University of Minnesota Human Research Protection Program (MMC 820

420 Delaware St. SE, Minneapolis, MN 55455-0392)

^*^Institution: University of Colorado Cancer Center

Name and address: Colorado Multiple Institutional Review Board (COMIRB) (University of Colorado, Anschutz Medical Campus, 13001 E. 17th Place, Building 500, Room N3214, Aurora, CO 80045)

^*^Institution: University of Washington Medical Center

Fred Hutchinson Cancer Research Center (FHCRC) IRB (Institutional Review Office 1100 Fairview Ave. N. Mail Stop J2-100, Seattle, WA 98109)

^*^Institution: University of Southern California, Norris Comprehensive Cancer Center University of Southern California Health Sciences Institutional Review Board (LAC+USC Medical Center, General Hospital Suite 4700, 1200 North State Street, Los Angeles, CA 90033)

^*^Institution: University of Texas, M.D. Anderson Cancer Center

University of Texas MD Anderson Cancer Ctr Clinical IRBs (Office of Human Subjects Protection

Unit 1637, 7007 Bertner Ave., Houston, TX 77030-3907)

^*^Institution: Swedish Cancer Institute

Western Institutional Review Board (WIRB) (1019 39th Avenue SE Suite 120, Puyallup, WA 98374-2115)

^*^Institution: University of Arizona, Arizona Cancer Center at UMC and UMC-North University of Arizona Institutional Review Board (The University of Chicago Biological Sciences Division/University of Chicago Medical Center, 5751 S. Woodlawn Ave., 2nd floor, Chicago, IL 60637)

All patients recruited in I-SPY2 signed an informed consent form.

The present study using the Kamilla samples was approved by the institutional research review board (Oslo University Hospital, Department of Cancer, Po Box 4953 Nydalen, 0424 Oslo), and the Regional Committees for Medical and Health Research Ethics (REC North-Secretariat, University of Tromo, Po Box 6050 Langnes, 9037 Tromsø). The patients still alive at the time of collection of the data presented here, signed a separate informed consent form.

## 3. RESULTS

### 3.1 The cellular efficacy of T-DM1 do not correlate to trastuzumab sensitivity

The antiproliferative effects of the HER2-targeted mAb trastuzumab and the intracellular acting HER2-targeted ADC T-DM1 were established in five HER2-positive cell lines. Subjecting the cells to a 72 hrs treatment with trastuzumab or T-DM1 revealed the SK- BR-3 and AU-565 cells as highly sensitive to both therapeutics, whereas the SKOV-3 cells, on the contrary, were found non-responsive to trastuzumab and exhibited low sensitivity to T-DM1 as demonstrated by a relatively high IC_50_ of 1.2 μg/ml (Fig. 1A and B). The HCC1954 and MDA-MB-453 cells were both found low- to moderately sensitive to trastuzumab treatment, but responded differently to T-DM1 with the HCC1954 cells showing high sensitivity, whereas the MDA-MB-453 cells showed low sensitivity (Fig. 1A and B). No clear connection was therefore found between trastuzumab and T-DM1 sensitivity among the five cell lines (Fig. 1A and B).

**Figure 1:**
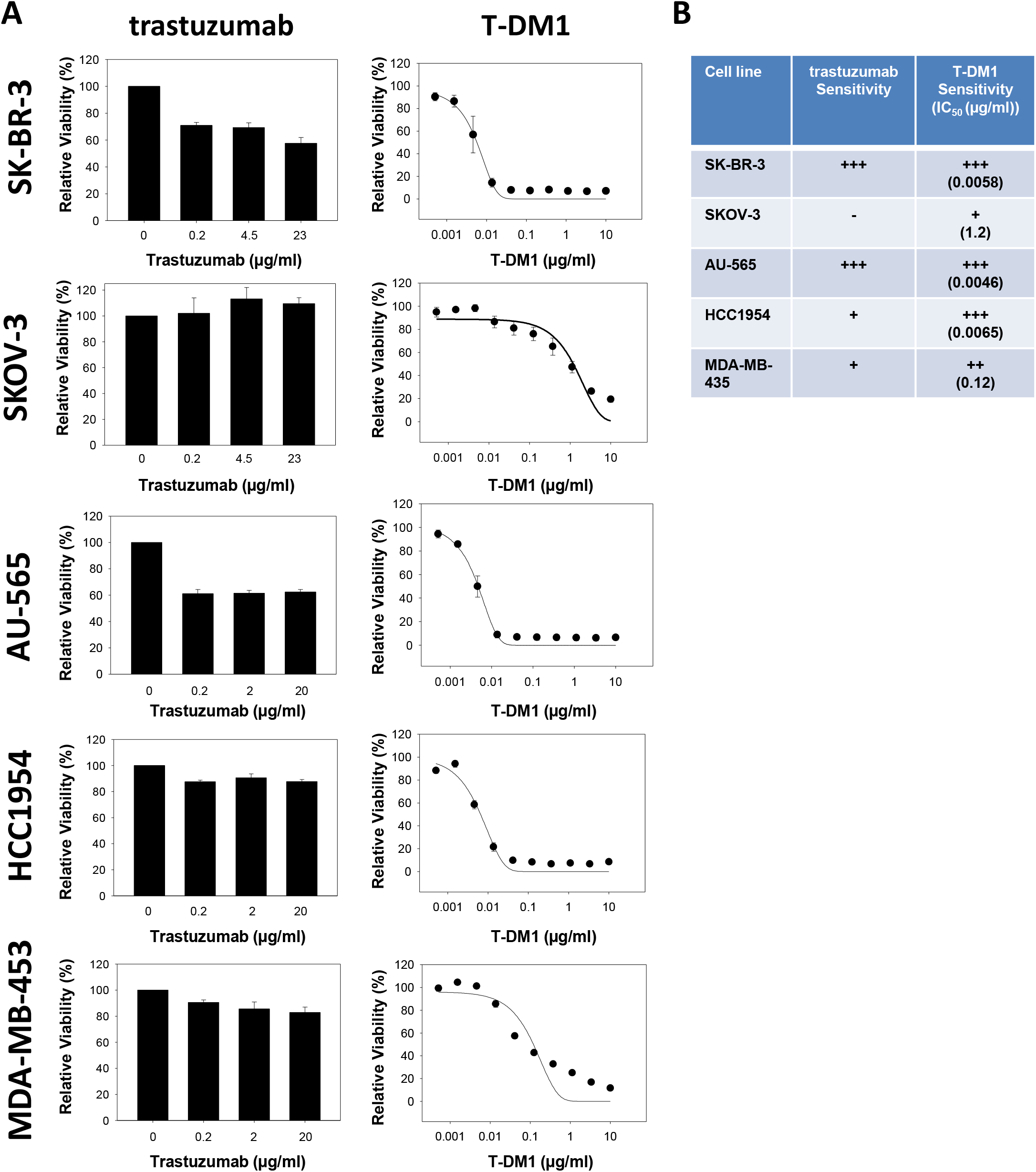
In vitro sensitivity to trastuzumab and T-DM1. A: Relative viability (MTT) of SK-BR-3, SKOV-3, AU-565, HCC1954 and MDA-MB-453 following 72 hrs treatments with indicated drugs. The sigmoid curve fit model a/(1 + exp(-(*x* - *x*_0_) / *b*)) was used for T-DM1. Data points represent the average of three independent experiments (trastuzumab, error bars: SE) or one representative of at least three separate experiments (T-DM1, error bars: SD). B: Cellular sensitivity of trastuzumab and T-DM1.

### 3.2 The T-DM1 sensitivity correlate to HER2 expression in HER2-positive breast and ovarian cancer cell lines

Strong HER2 expression was documented in the five HER2 expressing cell lines used in the present study compared to the low expression in MDA-MB-231 reported as HER2 negative (Fig. 2A)^26^.

**Figure 2:**
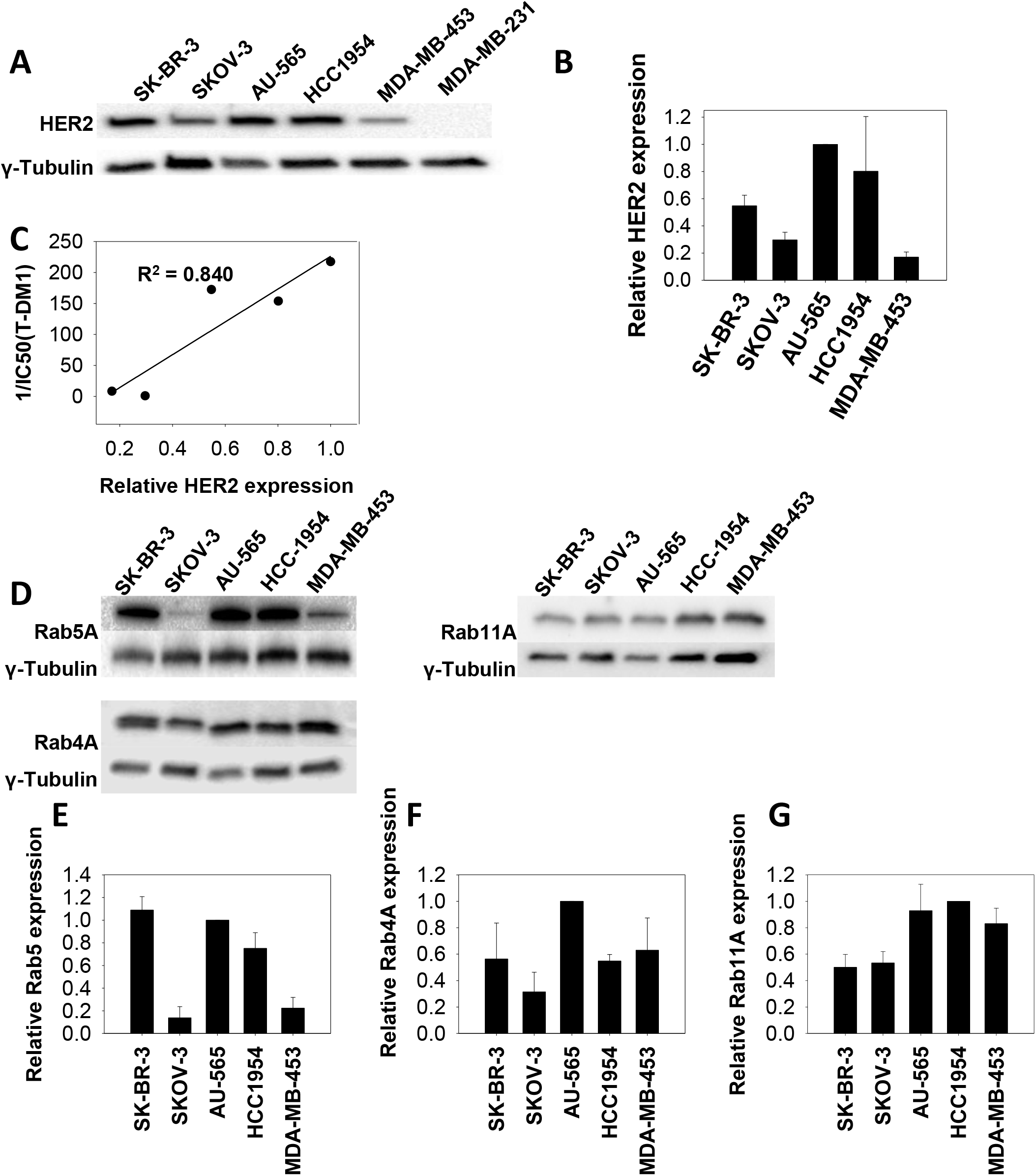
HER2 and RAB GTPase expression in cell lines. A: Western blot of HER2 and γ-tubulin expression in SK-BR-3, SKOV-3, HCC1954, AU-565, MDA-MB-453 and MDA-MB-231 cells. B shows quantification of the Western blot results based on 3 independent experiments (error bars: SD). C: Linear regression analysis curve of HER2 protein expression and and T-DM1 sensitivity (1/IC_50_(T-DM1)). Representative (D) and quantified (E-G) Western blots (n=3) of RAB4A, RAB5A, RAB11A and γ-tubulin expression in SK-BR-3, SKOV-3, AU-565, HCC1954 and MDA-MB-453 cells. Error bars: SD.

HER2 expression is essential for T-DM1 toxicity. However, it was hypothesized that the level of expression may not necessarily correlate directly to drug sensitivity due to differences in drug processing (e.g. uptake, intracellular transport and interaction with intracellular drug targets) between the cell lines. Quantification of HER2 expression in the five positive cell lines indicated AU-565 to have the highest expression level of HER2, closely followed by HCC1954 (Fig. 2A and B). A ∼50% lower HER2 expression was found in the SK-BR-3 cells compared to AU-565 while SKOV-3 and MDA-MB-453 were identified as the cell lines with the lowest HER2 expression in the panel (0.29 and 0.17 relative expression, respectively) (Fig 2A and B). The level of HER2 expression reported here is in agreement with other reports^26,27^. Furthermore, a linear relationship was found between HER2 expression and T-DM1 sensitivity among the cell lines, resulting in an R^2^ value of 0.840 (Fig. 2C).

### 3.3 HER2-expressing cell lines differ in their expression level of proteins involved in endocytic trafficking

As T-DM1 is dependent on internalization and intracellular trafficking in order to exert its intracellular mechanism of action, proteins essential for endocytosis and exocytosis were quantified in the cell line panel. These proteins included RAB5A (Fig. 2D and E), implicated in the delivery of cargo from the plasma membrane to early endosomes as well as endosome fusion, RAB4A (Fig. 2D and F), implicated in recycling from early endosomes and RAB11A (Fig 2D and G), involved in perinuclear recycling of endosomes and plasma membrane-Golgi traffic^28,29^. The expression level of these proteins among the cell lines showed large differences and no simple connection was found between the expression levels.

### 3.4 RAB5A protein expression is highly correlated to T-DM1 sensitivity

It was further assessed if the investigated proteins involved in endocytosis or exocytosis (Fig.2 D-G) had an impact on T-DM1 sensitivity in the evaluated cell lines. The expression level of RAB5A varied highly between the cell lines (Fig. 2D and E). However, a strong linear correlation was found between T-DM1 toxicity and RAB5A expression among the cell lines (R^2^ = 0.934) (Fig. 3A). In contrast, no linear correlations were found between T-DM1 sensitivity and expression of RAB4A or RAB11A (Fig. 3B and C). The cellular T-DM1 sensitivity was also correlated to the expression level of HER2 and RAB5A together revealing an R^2^ = 0.962 which is higher than obtained with the correlations of HER2 (R^2^ = 0.840) (Fig. 2C) and RAB5A (R^2^ = 0.934) (Fig. 3A) alone. Thus, T-DM1 sensitivity correlates to expression of RAB5A in this panel of cell lines, while no linear correlation is found for RAB4A and RAB11A. The expression level of HER2 and RAB5A together serves as a better biomarker for cellular T-DM1 response as compared to either of the two proteins alone.

**Figure 3:**
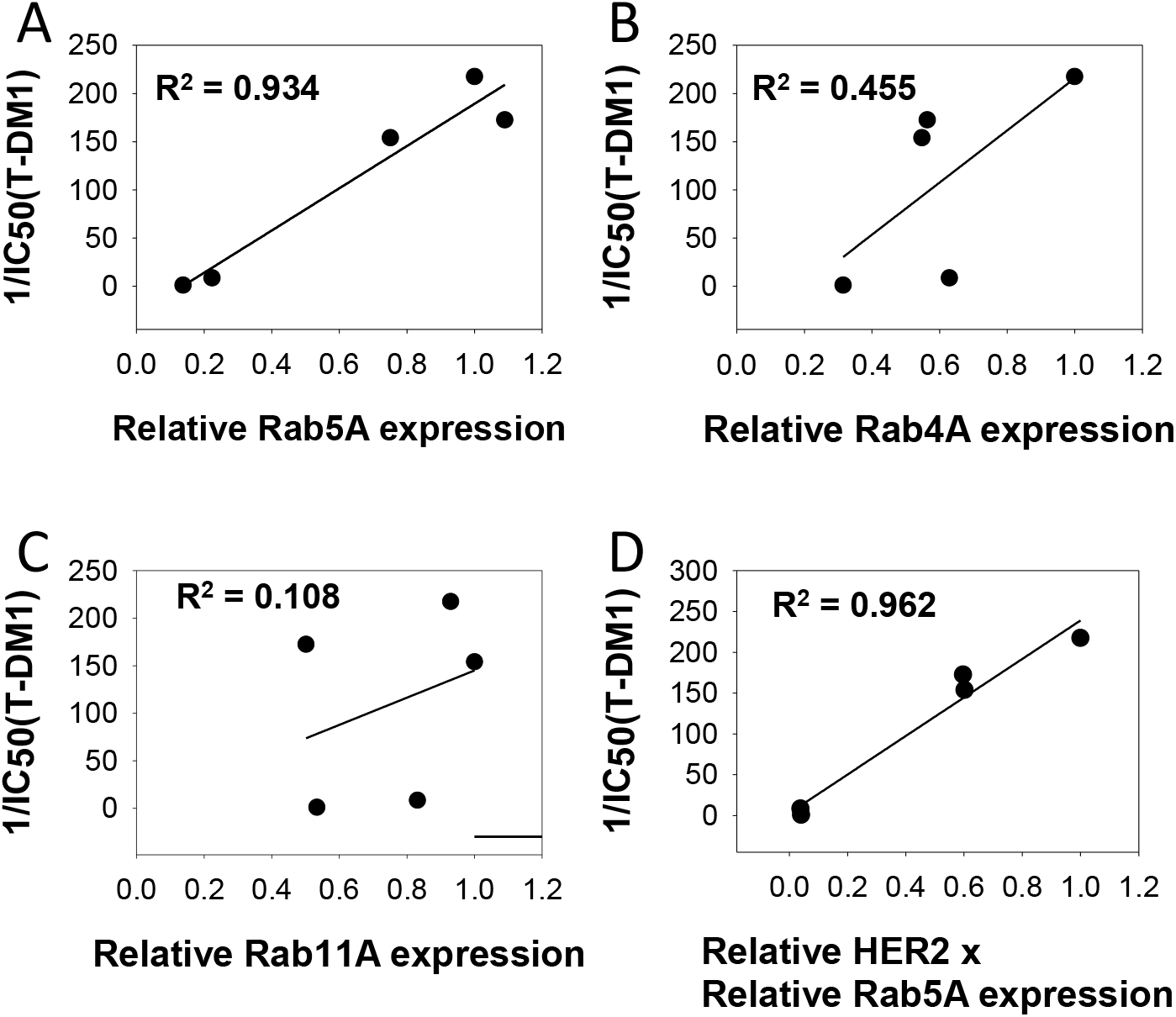
T-DM1 and RAB GTPase in vitro correlations. Linear regression analysis curves between RAB5A (A), RAB4A (B) and RAB11A (C) protein expression and T- DM1 sensitivity (1/IC_50_(T-DM1) in the five cell lines. D shows the linear regression curve between HER2 x RAB5A protein expression and T-DM1 response.

### 3.5 RAB5 RNA expression correlates with T-DM1 sensitivity in the I-SPY 2 clinical trial

We then validated our *in vitro* finding in the I-SPY 2 clinical trial by testing if the clinical response towards T-DM1, as measured by pCR, could be correlated to RAB5A RNA expression. Figure 4A summarizes the pCR and the hormone receptor (HR) status of the T-DM1 + pertuzumab (T-DM1+P) and trastuzumab and paclitaxel (TH) control arms. Overall, 30 of the 52 patients on the T-DM1+P arm and 8 of the 31 patients in the control arm achieved a pCR. Normalized, pre-treatment expression levels of RAB4A, RAB5A and RAB11A were tested individually as biomarkers of response. Of the biomarkers evaluated, only RAB5A was associated with response in the T-DM1 + pertuzumab arm (Fig.4B, p = 0.01, LR test). None of the tested biomarkers were associated with response in the control arm (trastuzumab + paclitaxel) (Fig. 4B). The p value for the interaction between RAB5A expression and treatment was 0.02 and remained < 0.05 after adjusting for hormone status (Fig. 4B).

**Figure 4:**
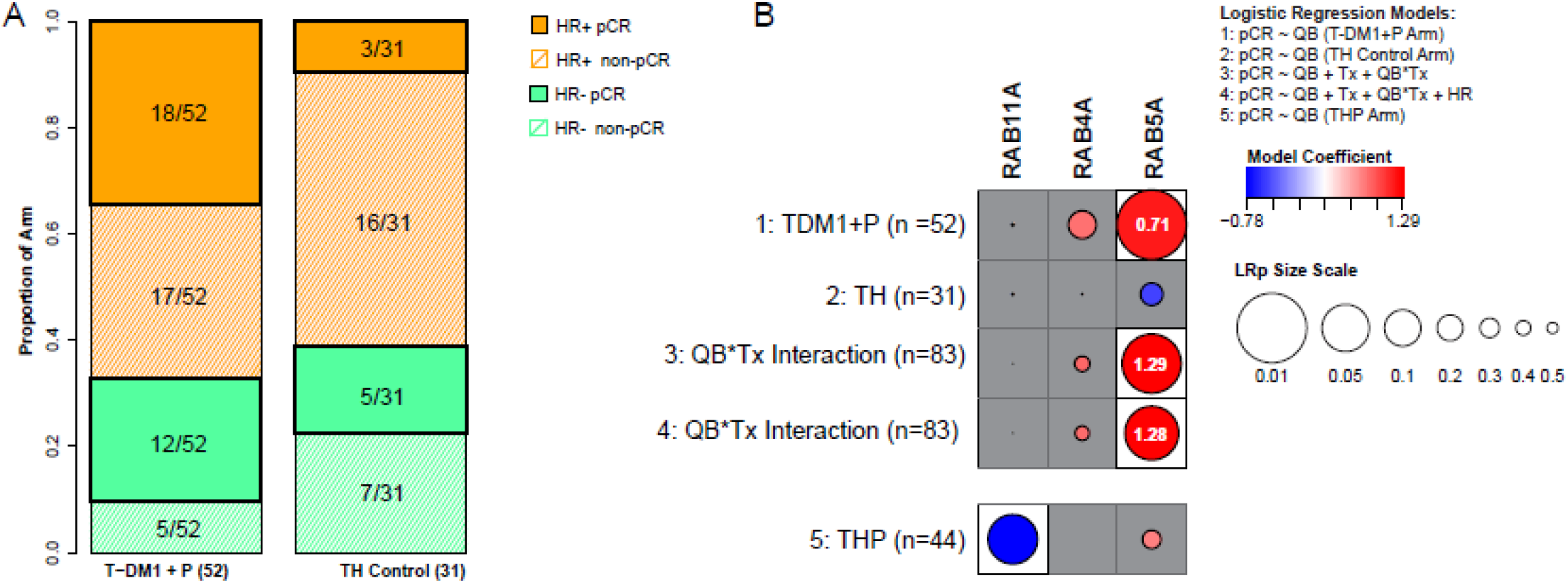
T-DM1 and RAB GTPase correlations in the I-SPY2 cohort. A: Number of patients, hormone receptor (HR) status and pathological complete responses (pCR) in the T-DM1+pertuzumab and trastuzumab paclitaxel arm of the I-SPY2 study. B: Association plot summarizing qualifying biomarker analyses of RAB4A, RAB5A and RAB11A expression levels as specific predictors of pCR to indicated treatment. Results are organized by the logistic model/data used along the rows; and the biomarker evaluated along the columns. Circle sizes are proportional to the significance (-log10 (LR test p)); and circle color reflect magnitude of coefficient (red: positive, blue: negative) from each corresponding logistic model. White background indicates p<0.05; and the odds ratio associated with 1 standard deviation increase in expression were also shown (in white) inside the circle.

The association between RAB5A and T-DM1+pertuzumab response may be attributable to pertuzumab rather than T-DM1. Although we cannot directly compare the two experimental arms, analysis of the trastuzumab+ paclitaxel+ pertuzumab group (n=44) showed no significant correlation between RAB5A and pCR (Coef = 0.38, LRp = 0.32) suggesting pertuzumab is not the key contributor to the significant pCR and RAB5A association for T-DM1+pertuzumab treated patients (Fig. 4B).

RAB5A is a constitutive expressed protein, and utilization as a biomarker will depend on the establishment of a threshold which separates RAB5A^high^ and RAB5A^low^ expressing cancer. Figure 5A shows boxplots of RAB5A expression levels stratified by arm and pCR status; based on a Monte-Carlo 2-fold cross validation procedure as described in M&M, a normalized RAB5A RNA expression level of 9.76 was selected as a threshold (dotted gray line). The patient stratification into RAB5A^high^ and RAB5A^low^ groups in each arm is illustrated in Fig. 5B together with the pCR data.

**Figure 5:**
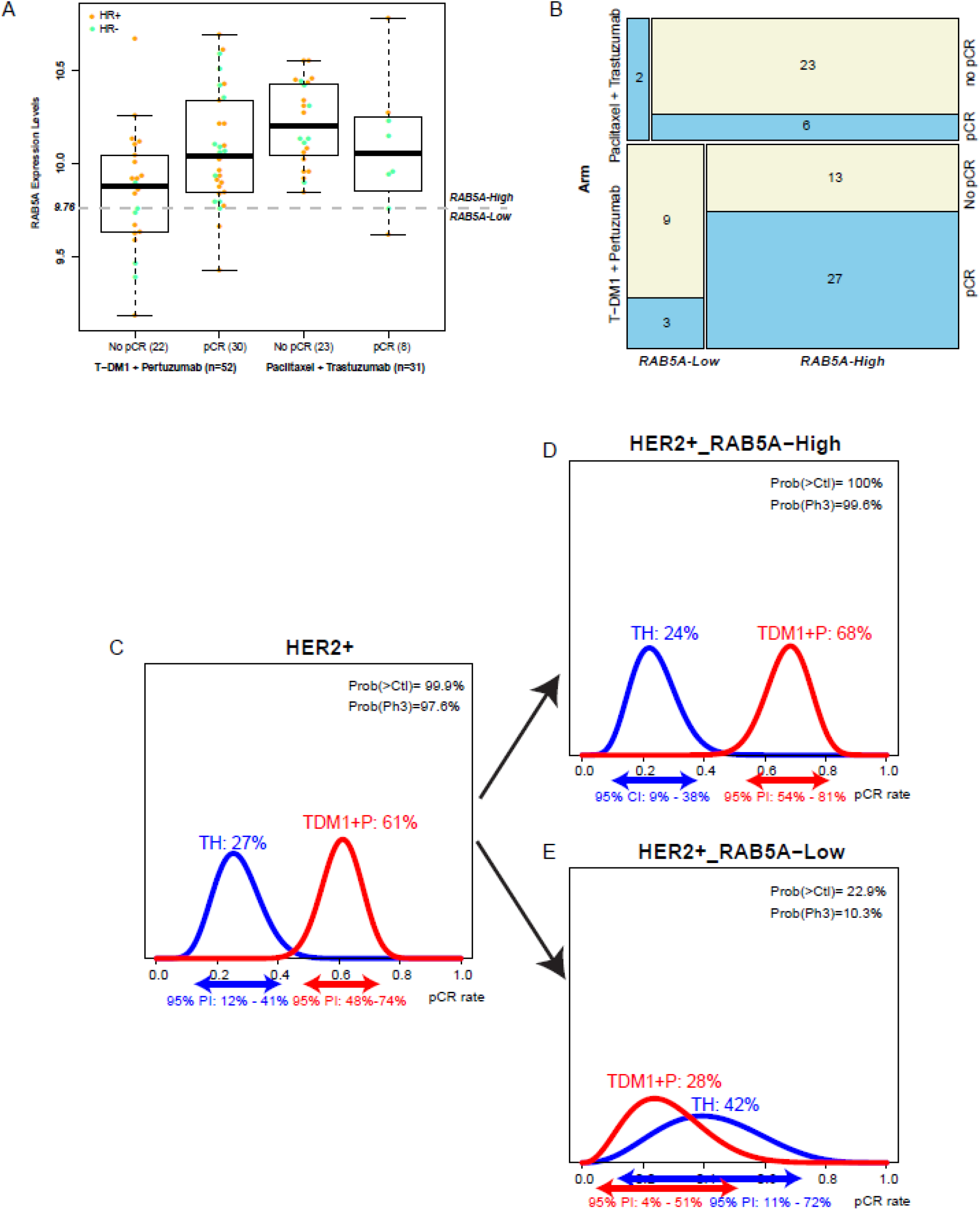
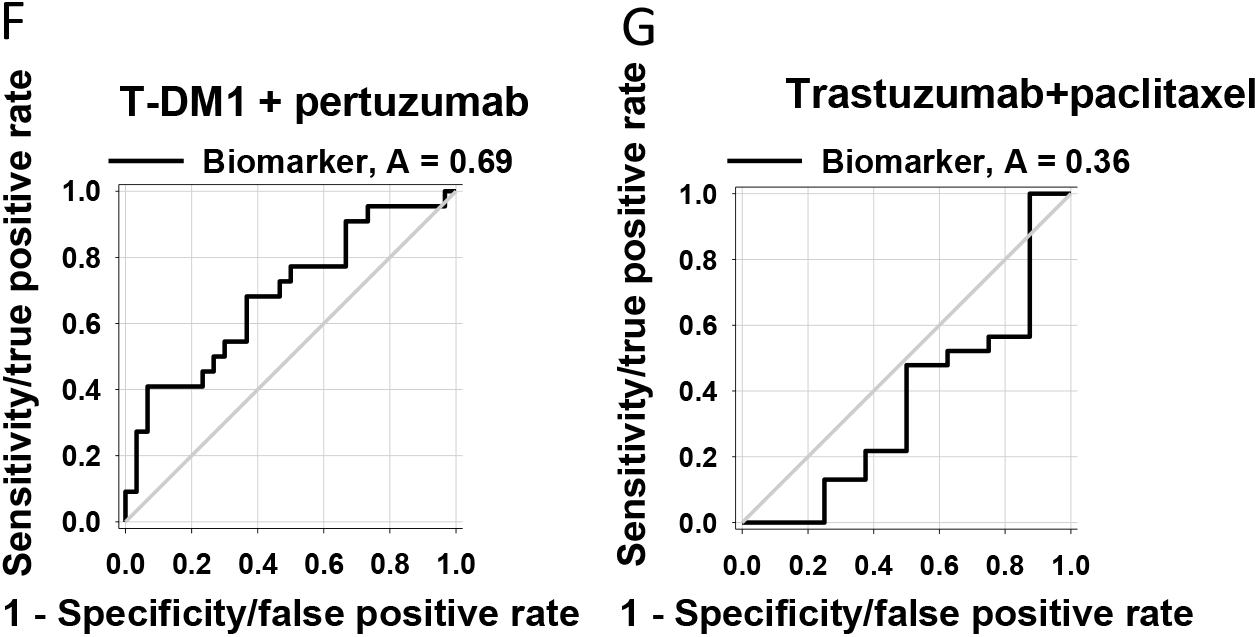
Patient distribution within PCR and RAB5A expression in I-SPY2. A: Box plot of RAB5A expression levels stratified by arm and pCR status. Dots represent expression values for each individual; and color reflects subtype (orange: HR+; green: HR-). B: Mosaic plot showing patient distribution within the RAB5A RNA-high and - low based on the threshold of 9.76 by arm and pCR status. C-E illustrates the bayesioan estimated pCR rates within the two treatment groups overall (C) as well as when divided in RAB5A RNA-high (D) and low (E) subsets. F and G shows ROC curves of the performance of RAB5A RNA as a biomarker in T-DM1 + pertuzumab treated (F)- and trastuzumab + paclitaxel treated (G) patients.

**Figure 6:**
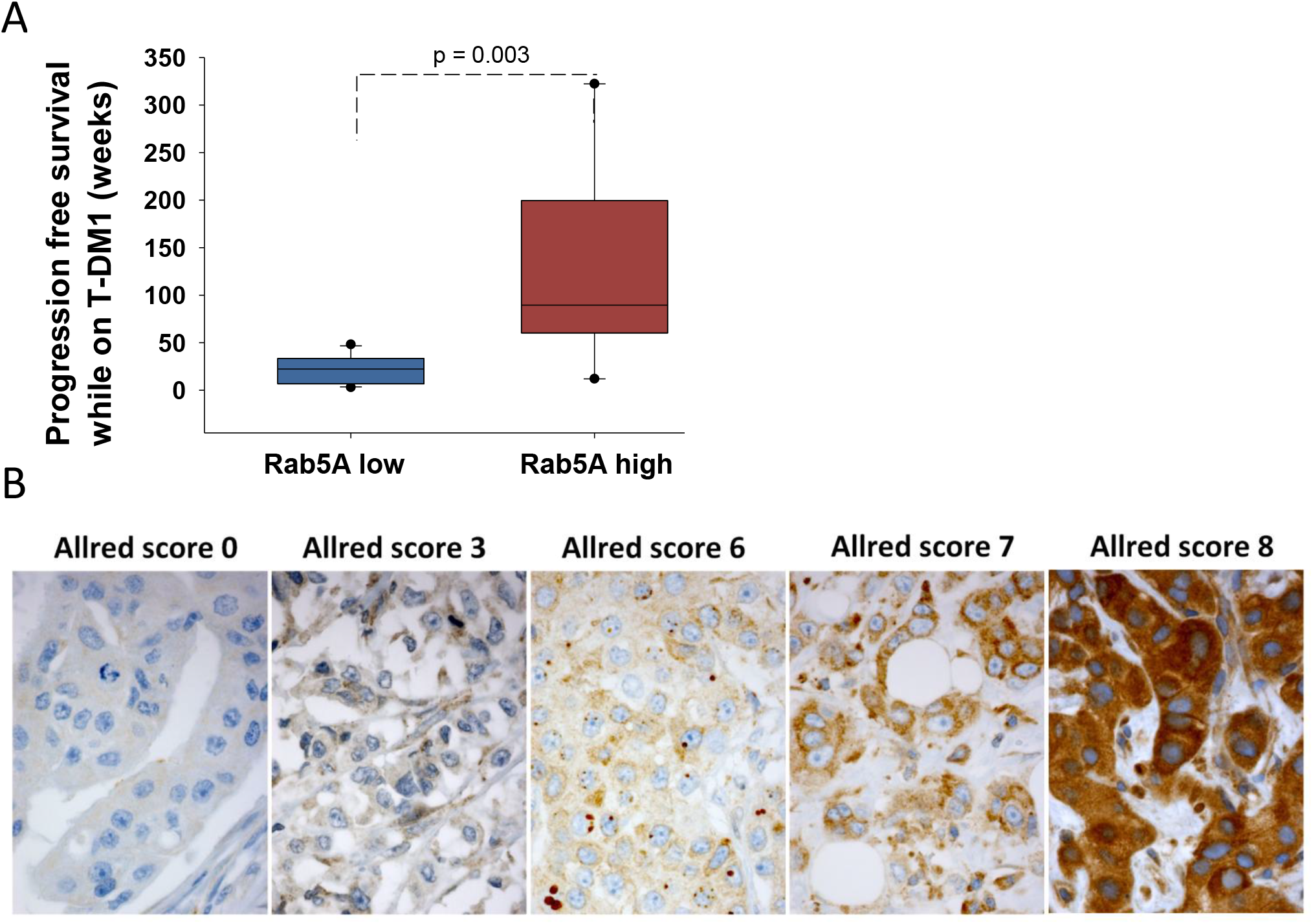
T-DM1 and RAB5A correlation in a subset of patients in KAMILLA. A: Box blot indicating the distribution of PFS in RAB5A low (Allred score 0-6) and RAB5A high (Allred score 7 and 8) expressing patients. B: Exemplified images (40x objective) of Rab5A IHC for Allred score 0 (patient #20) Allred score 3 (Patient # 19), Allred score 6 (patient #21), Allred score 7 (patient #3) and Allred score 8 (patient #15).

Using this optimal threshold, only 2 patients in the TH arm (6%) had RAB5A levels < 9.76 and are considered RAB5A^low^, as opposed to 23% (12/52) of the T-DM1+P arm (Fig. 5B). This may in part be attributed to the difference in RAB5A expression between the two arms, where the RAB5A levels are significantly higher in the trastuzumab+paclitaxel arm than the T-DM1+pertuzumab arm. Nevertheless, within the T-DM1+P arm, 68% (27/40) of RAB5A^high^ patients achieved a pCR, in contrast to 25% (3/12) of RAB5A^low^ patients (Fig. 5B). Of note, no significant differences in HR status distribution or pre-treatment ERBB2 expression levels are observed among the RAB5A^high^ and RAB5A^low^ T-DM1+P treated patients (% HR-: 30% vs. 42%, Fisher exact test p=0.49, and median ERBB2: 11.2 vs. 10.6, Wilcoxon rank sum p=0.08 respectively).

Bayesian logistic modelling was used to estimate the pCR probability distributions within the T-DM1+P and TH control arms in the overall HER2+ population as well as within the (predicted sensitive) RAB5A^high^ and (predicted insensitive) RAB5A^low^ subsets (Fig5C-E). The Bayesian estimated pCR probability is 68% in the T-DM1+pertuzumab arm relative to 24% in the control arm in the RAB5A^high^ patients. In contrast, the estimated pCR probability is 28% in the RAB5A^low^ subset in the T-DM1+pertuzumab arm and 42% in the trastuzumab+paclitaxel arm. For comparison, using the same model, the estimated pCR probability of the entire HER2+ group is 61% in the TDM1+pertuzumab arm and 27% in the trastuzumab+paclitaxel arm.

Finally we also established ROC curves to find the most appropriate cut off for RAB5A expression in the T-DM1 + pertuzumab arm. Using this method, the optimal RAB5A cut off was found at 9.76, the same as the one identified using the Monte-Carlo procedure. The AUC was 0.69, again indicating RAB5A as a predictive biomarker in this cohort (Fig. 5F). A ROC curve was also calculated for the TH control arm. (Fig. 5G). In contrast to the T-DM1+P arm, the low AUC of 0.36 (below 0.5) suggested low RAB5A expression as a potential predictor for trastuzumab+paclitaxel response, although the number of patients (n =2) with RAB5A^low^ in this arm is too small to evaluate (Fig. 5G).

### 3.6 RAB5A protein expression correlates with T-DM1 sensitivity in a subset of patients in the Kamilla study

Our positive correlation between RAB5A expression and T-DM1 sensitivity both *in vitro* and in the I-SPY2 clinical cohort was further verified in a small subset of the KAMILLA study including 19 patients treated at Oslo University hospital with biobanked primary biopsies. A highly significant (p< 0.03) difference in PFS was found between patients with RAB5A Allred scores 7 and 8, where 7/8 patients had a PFS of 50 weeks or more, and Allred scores 0-6, where all patients had PFS less than 50 weeks (6A). The Allred score and PFS data for each patients is included in Supplementary table 2 and exemplified IHC images are shown in 6B.

## DISCUSSION

Most of the targeting drugs currently approved for the treatment of cancer are mAbs or small-molecular inhibitors for which the drug target also represents the target for mechanism of action. The target itself represents a clear biomarker for treatment with these drugs, although other factors may be important to identify patients likely to experience resistance or low tolerability. In complex targeting therapeutics incorporating a cytotoxic component such as in ADCs, other biomarkers associated with the intracellular transport and/or cytotoxic mechanism of action are likely to impact on the therapeutic outcome^12,13^. In this study, the need for separate predictive biomarkers for ADC and mAbs treatment is emphasized by the lack of coherence between trastuzumab and T-DM1 sensitivity in the selected panel of HER2-positive cell lines. We demonstrate a linear correlation between cellular HER2 expression and response towards T-DM1 (Fig. 2C). This is in agreement with several clinical studies demonstrating higher response rates of T-DM1 in patients with HER2 mRNA levels above the median compared to the below median subgroup^14,15,30^. Importantly, despite its highly targeted mechanism, T-DM1 shows clinical benefit only in a subset of HER2- positive breast cancer patients with an objective response rate reported to ∼40% ^8,31,32^. Furthermore, in HER2 positive gastric cancer an objective T-DM1 response rate of only ∼20% with no increase in efficacy compared to taxanes is reported^33^. The diverse clinical response towards T-DM1 in HER2 expressing cancer indicate drug efficacy to depend also on other factors than the extracellular target expression.

T-DM1 is dependent on endocytosis in order to transport its cytotoxic payload (emtansine) into the cell and we here present the first report on T-DM1 efficacy correlated to proteins involved in endocytosis and endocytic trafficking. Of the 3 proteins investigated (RAB4A, RAB5A and RAB11A), only RAB5 expression was found to correlate with T-DM1 toxicity in a cell line panel of HER2-positive breast and ovarian cancer. The correlation of T-DM1 sensitivity and RAB5A protein expression was stronger than observed for HER2 expression, and the highest correlation was found when combining RAB5A and HER2 protein expression indicating incorporation of RAB5A together with HER2 as a better predictive biomarker for T-DM1 sensitivity. RAB5A is localized to early endosomes and regulates both endocytosis and endosome fusion of clathrin-coated vesicles^34^. The lack of correlation observed for RAB4A and RAB11A may reflect the intracellular processing of T-DM1 upon uptake. RAB4A mediated recycling from early endosomes may e.g. be limited ^35-37^ and it is possible that DM1 escape the endocytic vesicles prior to accumulation in RAB11A-positive recycling endosomes. We confirmed the correlation between RAB5A expression and sensitivity to neoadjuvant T-DM1 in the I-SPY2 study. Even though the number of patients is small, our clinical data from I-SPY2 also illustrates the possibility to define a RAB5A expression threshold to determine T-DM1 treatment in HER2 expressing breast cancer patients. A subset of patients from the Kamilla study was here used as a verification cohort. As compared to the I-SPY2 cohort, the patients from the Kamilla cohort all had advanced progressive disease and had all previously been exposed to different treatment regimens, including chemo- and HER2- targeted therapies. Nevertheless, a highly significant correlation was found between RAB5A expression in primary tumor and PFS while on T-DM1 therapy. Altogether, our presented *in vitro* findings showing a significant correlation between T-DM1 treatment outcome and RAB5A expression, is here confirmed in two independent clinical cohorts.

It has previously been shown that RAB5A expression predict poor prognosis of breast cancer patients^38^, and the current study strongly indicate that T-DM1 should be further evaluated as the treatment of choice for these patients.

T-DM1 is currently approved only for advanced HER2-positive breast cancer previously treated with trastuzumab, but a defined RAB5A threshold-biomarker holds promise also for patient stratification at earlier stages of this disease^39^, in addition to other indications, such as gastric cancer^33^.

Although the number of patients with RAB5A^low^ receiving trastuzumab in the I-SPY2 cohort is far too low to draw any conclusions, our results may also point towards a negative correlation between RAB5A expression and trastuzumab response. Thus, HER2 expressing cancers with low RAB5A do better on trastuzumab as compared to T- DM1. If low RAB5A can be used not only to deselect patients for T-DM1 but also to find those patients most likely to benefit from trastuzumab, it would be an improvement for future personalized HER2 positive breast cancer therapy. Altogether, better stratification of HER2 expressing breast cancer patients into treatment groups may add to the benefit of the treatment, and the deselected patients may also be offered alternative treatment at an earlier time point.

## Conclusion

As the mechanism of T-DM1 action involves endocytosis, we hypothesized that proteins involved in the endocytic process had impact on the treatment response to T-DM1. Our hypothesis was here confirmed first *in vitro* and then in two independent clinical cohorts and the present report, for the first time, shows that RAB5A may be used to predict the response to T-DM1. Furthermore we believe our results also demonstrate a more general novel concept in which proteins involved in endocytosis and/or endocytic trafficking is utilized as biomarkers for ADCs. Even though RAB5A was the only candidate to succeed as a predictive biomarker for T-DM1 in the present study, this may be different for other ADCs dependent on both their targeting moiety and cytotoxic payload. Overall, our results imply that current and future ADC development and treatment will benefit from incorporation of biomarkers reflecting uptake and intracellular transport.

## Data Availability

The clinical data referred to in the manuscript is available in the supplementary file

