## Supplemental Table 1 and 2 for "RAB5A expression is a predictive biomarker for trastuzumab emtansine in breast cancer"

### Supplementary Figures

**Supplementary Table 1**
**Normalized, platform-corrected pre-treatment expression levels of RAB5A, RAB4A and RAB11A from patients in the indicated treatment arms. The data are blinded.**

| ResearchID | Arm | HR | HER2 | pCR | RAB11A | RAB4A | RAB5A |
|---|---|---|---|---|---|---|---|
| 146033 | Paclitaxel + Pertuzumab + Trastuzumab | 1 | 1 | 1 | 9.9596 | 9.5662 | 10.098 |
| 551795 | Paclitaxel + Pertuzumab + Trastuzumab | 0 | 1 | 1 | 9.5884 | 10.247 | 10.124 |
| 570685 | Paclitaxel + Pertuzumab + Trastuzumab | 1 | 1 | 0 | 10.182 | 10.694 | 9.9122 |
| 999264 | Paclitaxel + Pertuzumab + Trastuzumab | 0 | 1 | 1 | 8.431 | 9.4374 | 10.183 |
| 960029 | Paclitaxel + Pertuzumab + Trastuzumab | 1 | 1 | 0 | 10.288 | 9.7121 | 10.605 |
| 618717 | Paclitaxel + Pertuzumab + Trastuzumab | 1 | 1 | 0 | 9.8212 | 8.7855 | 9.7853 |
| 795996 | Paclitaxel + Pertuzumab + Trastuzumab | 1 | 1 | 1 | 9.9511 | 9.7984 | 10.241 |
| 981664 | Paclitaxel + Pertuzumab + Trastuzumab | 1 | 1 | 1 | 9.637 | 9.6174 | 10.08 |
| 285339 | Paclitaxel + Pertuzumab + Trastuzumab | 0 | 1 | 1 | 9.6838 | 8.8935 | 10.07 |
| 483667 | Paclitaxel + Pertuzumab + Trastuzumab | 1 | 1 | 0 | 10.192 | 8.5211 | 10.403 |
| 432114 | Paclitaxel + Pertuzumab + Trastuzumab | 1 | 1 | 1 | 10.045 | 9.5937 | 10.113 |
| 474874 | Paclitaxel + Pertuzumab + Trastuzumab | 0 | 1 | 1 | 9.6295 | 8.9655 | 9.7843 |
| 273691 | Paclitaxel + Pertuzumab + Trastuzumab | 1 | 1 | 1 | 10.065 | 10.318 | 10.074 |
| 527581 | Paclitaxel + Pertuzumab + Trastuzumab | 1 | 1 | 0 | 9.0901 | 9.5776 | 9.9122 |
| 738041 | Paclitaxel + Pertuzumab + Trastuzumab | 1 | 1 | 1 | 10.188 | 9.4336 | 10.552 |
| 900959 | Paclitaxel + Pertuzumab + Trastuzumab | 1 | 1 | 0 | 9.9455 | 8.6225 | 9.8532 |
| 991818 | Paclitaxel + Pertuzumab + Trastuzumab | 1 | 1 | 1 | 9.8885 | 9.3862 | 10.121 |
| 575457 | Paclitaxel + Pertuzumab + Trastuzumab | 0 | 1 | 1 | 9.3164 | 9.6164 | 10.465 |
| 296841 | Paclitaxel + Pertuzumab + Trastuzumab | 1 | 1 | 1 | 9.6426 | 10.084 | 10.342 |
| 982086 | Paclitaxel + Pertuzumab + Trastuzumab | 0 | 1 | 1 | 9.2575 | 10.526 | 10.508 |
| 721099 | Paclitaxel + Pertuzumab + Trastuzumab | 1 | 1 | 0 | 9.4669 | 10.415 | 9.5461 |
| 453596 | Paclitaxel + Pertuzumab + Trastuzumab | 1 | 1 | 1 | 9.9474 | 9.2109 | 10.377 |
| 825363 | Paclitaxel + Pertuzumab + Trastuzumab | 0 | 1 | 1 | 9.6408 | 9.3047 | 10.331 |
| 862244 | Paclitaxel + Pertuzumab + Trastuzumab | 0 | 1 | 0 | 9.0649 | 9.5113 | 10.286 |
| 377460 | Paclitaxel + Pertuzumab + Trastuzumab | 0 | 1 | 1 | 8.7938 | 9.7586 | 10.03 |
| 144302 | Paclitaxel + Pertuzumab + Trastuzumab | 1 | 1 | 0 | 9.7716 | 8.307 | 9.8935 |
| 462605 | Paclitaxel + Pertuzumab + Trastuzumab | 0 | 1 | 1 | 9.3089 | 10.275 | 10.003 |
| 204939 | Paclitaxel + Pertuzumab + Trastuzumab | 1 | 1 | 1 | 9.8408 | 9.0138 | 10.177 |
| 472799 | Paclitaxel + Pertuzumab + Trastuzumab | 0 | 1 | 1 | 9.5744 | 9.2138 | 10.131 |
| 685182 | Paclitaxel + Pertuzumab + Trastuzumab | 1 | 1 | 1 | 9.8259 | 9.1493 | 10.239 |
| 649198 | Paclitaxel + Pertuzumab + Trastuzumab | 0 | 1 | 0 | 9.6295 | 9.8884 | 10.389 |
| 127791 | Paclitaxel + Pertuzumab + Trastuzumab | 1 | 1 | 0 | 10.258 | 10.254 | 9.7715 |
| 235995 | Paclitaxel + Pertuzumab + Trastuzumab | 1 | 1 | 1 | 9.394 | 8.3165 | 9.8807 |
| 203060 | Paclitaxel + Pertuzumab + Trastuzumab | 1 | 1 | 0 | 10.037 | 9.8903 | 10.294 |
| 604326 | Paclitaxel + Pertuzumab + Trastuzumab | 1 | 1 | 0 | 9.8464 | 9.6363 | 10.364 |
| 982505 | Paclitaxel + Pertuzumab + Trastuzumab | 1 | 1 | 0 | 10.135 | 9.0347 | 10.176 |
| 799149 | Paclitaxel + Pertuzumab + Trastuzumab | 1 | 1 | 1 | 9.4903 | 9.4743 | 10.047 |
| 324021 | Paclitaxel + Pertuzumab + Trastuzumab | 1 | 1 | 0 | 9.7492 | 8.8442 | 9.7242 |

| | | | | | | | |
|---|---|---|---|---|---|---|---|
| 644609 | Paclitaxel + Pertuzumab + Trastuzumab | 0 | 1 | 1 | 9.594 | 9.3578 | 9.8896 |
| 895653 | Paclitaxel + Pertuzumab + Trastuzumab | 1 | 1 | 1 | 8.7143 | 8.8622 | 9.9821 |
| 503034 | Paclitaxel + Pertuzumab + Trastuzumab | 0 | 1 | 0 | 9.1574 | 9.4279 | 9.9319 |
| 995480 | Paclitaxel + Pertuzumab + Trastuzumab | 1 | 1 | 0 | 9.7492 | 9.8363 | 9.8925 |
| 584561 | Paclitaxel + Pertuzumab + Trastuzumab | 0 | 1 | 1 | 9.4772 | 10.293 | 9.8758 |
| 507249 | Paclitaxel + Pertuzumab + Trastuzumab | 1 | 1 | 0 | 10.585 | 10.537 | 10.486 |
| 402265 | Paclitaxel + Trastuzumab | 1 | 1 | 0 | 9.4237 | 9.5886 | 10.082 |
| 246134 | Paclitaxel + Trastuzumab | 1 | 1 | 0 | 9.5615 | 8.9417 | 10.342 |
| 220471 | Paclitaxel + Trastuzumab | 0 | 1 | 0 | 10.188 | 9.914 | 10.444 |
| 266840 | Paclitaxel + Trastuzumab | 1 | 1 | 0 | 10.109 | 9.8192 | 10.554 |
| 166412 | Paclitaxel + Trastuzumab | 1 | 1 | 0 | 10.057 | 8.3756 | 10.204 |
| 501303 | Paclitaxel + Trastuzumab | 0 | 1 | 1 | 9.4744 | 9.5966 | 10.231 |
| 141782 | Paclitaxel + Trastuzumab | 1 | 1 | 0 | 9.3724 | 9.665 | 9.8482 |
| 906795 | Paclitaxel + Trastuzumab | 1 | 1 | 0 | 10.062 | 10.211 | 10.025 |
| 375455 | Paclitaxel + Trastuzumab | 1 | 1 | 0 | 8.8622 | 9.8597 | 10.308 |
| 153880 | Paclitaxel + Trastuzumab | 1 | 1 | 0 | 10.354 | 10.068 | 10.273 |
| 539904 | Paclitaxel + Trastuzumab | 1 | 1 | 0 | 10.176 | 9.4696 | 10.438 |
| 596079 | Paclitaxel + Trastuzumab | 0 | 1 | 1 | 9.339 | 9.0599 | 10.149 |
| 856159 | Paclitaxel + Trastuzumab | 1 | 1 | 0 | 10.29 | 9.5255 | 10.452 |
| 797427 | Paclitaxel + Trastuzumab | 1 | 1 | 0 | 9.8567 | 10.505 | 9.9234 |
| 809649 | Paclitaxel + Trastuzumab | 0 | 1 | 1 | 9.3217 | 10.113 | 9.7566 |
| 889049 | Paclitaxel + Trastuzumab | 0 | 1 | 0 | 9.644 | 10.103 | 10.309 |
| 335091 | Paclitaxel + Trastuzumab | 1 | 1 | 1 | 10.92 | 10.055 | 10.781 |
| 382853 | Paclitaxel + Trastuzumab | 0 | 1 | 1 | 9.7795 | 9.5078 | 9.959 |
| 627118 | Paclitaxel + Trastuzumab | 0 | 1 | 0 | 8.838 | 10.474 | 9.8961 |
| 350331 | Paclitaxel + Trastuzumab | 0 | 1 | 0 | 9.4393 | 9.182 | 10.423 |
| 573586 | Paclitaxel + Trastuzumab | 1 | 1 | 0 | 10.272 | 9.1441 | 10.551 |
| 725455 | Paclitaxel + Trastuzumab | 0 | 1 | 0 | 8.8818 | 10.117 | 10.131 |
| 123660 | Paclitaxel + Trastuzumab | 0 | 1 | 0 | 10.322 | 9.8116 | 10.131 |
| 859699 | Paclitaxel + Trastuzumab | 1 | 1 | 1 | 9.3096 | 8.8865 | 10.276 |
| 824234 | Paclitaxel + Trastuzumab | 1 | 1 | 1 | 9.724 | 9.7766 | 9.6199 |
| 726381 | Paclitaxel + Trastuzumab | 1 | 1 | 0 | 9.6348 | 9.1759 | 9.9533 |
| 442637 | Paclitaxel + Trastuzumab | 0 | 1 | 0 | 8.8703 | 9.5869 | 10.115 |
| 742477 | Paclitaxel + Trastuzumab | 1 | 1 | 0 | 10.177 | 10.25 | 10.061 |
| 162562 | Paclitaxel + Trastuzumab | 0 | 1 | 1 | 9.9502 | 10.004 | 9.9418 |
| 720065 | Paclitaxel + Trastuzumab | 1 | 1 | 0 | 9.456 | 9.6316 | 9.9585 |
| 112606 | Paclitaxel + Trastuzumab | 1 | 1 | 0 | 9.143 | 8.3354 | 10.456 |
| 366120 | T-DM1 + Pertuzumab | 1 | 1 | 0 | 9.2808 | 9.028 | 9.1819 |
| 767081 | T-DM1 + Pertuzumab | 0 | 1 | 1 | 10.226 | 10.211 | 10.42 |
| 464547 | T-DM1 + Pertuzumab | 1 | 1 | 1 | 9.7829 | 9.5823 | 9.9359 |
| 220925 | T-DM1 + Pertuzumab | 0 | 1 | 0 | 8.9705 | 9.7567 | 9.8965 |
| 167271 | T-DM1 + Pertuzumab | 1 | 1 | 0 | 10.091 | 9.5899 | 10.044 |
| 947398 | T-DM1 + Pertuzumab | 1 | 1 | 1 | 10.279 | 9.3057 | 10.692 |
| 413211 | T-DM1 + Pertuzumab | 1 | 1 | 0 | 10.009 | 9.2915 | 10.258 |
| 786003 | T-DM1 + Pertuzumab | 1 | 1 | 1 | 9.5435 | 9.0811 | 9.8994 |

| | | | | | | | |
|---|---|---|---|---|---|---|---|
| 494022 | T-DM1 + Pertuzumab | 0 | 1 | 0 | 9.3986 | 10.618 | 9.4614 |
| 131961 | T-DM1 + Pertuzumab | 1 | 1 | 1 | 9.8362 | 10.205 | 10.613 |
| 208650 | T-DM1 + Pertuzumab | 1 | 1 | 0 | 10.285 | 9.7065 | 9.5904 |
| 490805 | T-DM1 + Pertuzumab | 1 | 1 | 1 | 9.6137 | 9.6733 | 9.9654 |
| 250274 | T-DM1 + Pertuzumab | 1 | 1 | 0 | 9.7352 | 9.4269 | 9.9329 |
| 410083 | T-DM1 + Pertuzumab | 0 | 1 | 1 | 9.8474 | 9.768 | 10.066 |
| 253531 | T-DM1 + Pertuzumab | 1 | 1 | 1 | 10.136 | 10.497 | 10.213 |
| 122287 | T-DM1 + Pertuzumab | 1 | 1 | 1 | 8.5629 | 8.7049 | 9.6652 |
| 491779 | T-DM1 + Pertuzumab | 1 | 1 | 1 | 9.4323 | 9.6581 | 9.8748 |
| 138425 | T-DM1 + Pertuzumab | 0 | 1 | 1 | 9.3313 | 10.113 | 10.508 |
| 831848 | T-DM1 + Pertuzumab | 0 | 1 | 1 | 9.7614 | 9.1105 | 10.355 |
| 356431 | T-DM1 + Pertuzumab | 0 | 1 | 1 | 9.8735 | 9.679 | 9.9368 |
| 329941 | T-DM1 + Pertuzumab | 1 | 1 | 1 | 9.5323 | 9.2526 | 10.218 |
| 831702 | T-DM1 + Pertuzumab | 1 | 1 | 0 | 8.6797 | 8.9333 | 9.9211 |
| 397505 | T-DM1 + Pertuzumab | 1 | 1 | 1 | 9.6651 | 9.1304 | 10.339 |
| 268937 | T-DM1 + Pertuzumab | 1 | 1 | 1 | 10.574 | 9.8789 | 9.8463 |
| 896852 | T-DM1 + Pertuzumab | 1 | 1 | 0 | 10.09 | 9.0025 | 10.108 |
| 535569 | T-DM1 + Pertuzumab | 0 | 1 | 0 | 8.9817 | 9.1626 | 9.7567 |
| 797499 | T-DM1 + Pertuzumab | 0 | 1 | 1 | 9.5865 | 9.5189 | 10.101 |
| 760011 | T-DM1 + Pertuzumab | 1 | 1 | 1 | 8.9134 | 9.1181 | 9.8374 |
| 125130 | T-DM1 + Pertuzumab | 0 | 1 | 1 | 9.1406 | 9.7368 | 9.7941 |
| 254273 | T-DM1 + Pertuzumab | 1 | 1 | 0 | 9.3379 | 9.8391 | 9.864 |
| 598835 | T-DM1 + Pertuzumab | 0 | 1 | 0 | 8.9564 | 9.4791 | 9.3866 |
| 953885 | T-DM1 + Pertuzumab | 1 | 1 | 0 | 9.1584 | 8.2985 | 9.6229 |
| 437162 | T-DM1 + Pertuzumab | 1 | 1 | 0 | 10.808 | 9.1512 | 10.669 |
| 536253 | T-DM1 + Pertuzumab | 1 | 1 | 1 | 9.9736 | 10.027 | 9.7695 |
| 213913 | T-DM1 + Pertuzumab | 0 | 1 | 1 | 9.5791 | 9.4317 | 10.093 |
| 587691 | T-DM1 + Pertuzumab | 1 | 1 | 1 | 9.4931 | 9.4431 | 10.429 |
| 624777 | T-DM1 + Pertuzumab | 1 | 1 | 0 | 10.257 | 9.2081 | 9.6662 |
| 824350 | T-DM1 + Pertuzumab | 0 | 1 | 1 | 9.5351 | 9.2877 | 10.595 |
| 705381 | T-DM1 + Pertuzumab | 1 | 1 | 1 | 10.79 | 9.1844 | 10.094 |
| 981461 | T-DM1 + Pertuzumab | 1 | 1 | 0 | 9.881 | 8.9115 | 9.9231 |
| 396723 | T-DM1 + Pertuzumab | 1 | 1 | 0 | 9.6856 | 9.7046 | 10.131 |
| 845079 | T-DM1 + Pertuzumab | 1 | 1 | 1 | 9.9147 | 9.1635 | 9.9132 |
| 706418 | T-DM1 + Pertuzumab | 0 | 1 | 1 | 8.7508 | 8.7305 | 9.7961 |
| 727255 | T-DM1 + Pertuzumab | 0 | 1 | 0 | 8.576 | 9.6657 | 9.7321 |
| 790397 | T-DM1 + Pertuzumab | 1 | 1 | 1 | 9.7445 | 9.7946 | 9.428 |
| 311316 | T-DM1 + Pertuzumab | 1 | 1 | 0 | 9.9717 | 9.1408 | 9.6288 |
| 889721 | T-DM1 + Pertuzumab | 0 | 1 | 1 | 9.1799 | 9.732 | 9.7597 |
| 865416 | T-DM1 + Pertuzumab | 1 | 1 | 0 | 9.723 | 9.5984 | 10.011 |
| 128051 | T-DM1 + Pertuzumab | 1 | 1 | 0 | 10.411 | 8.9466 | 9.8423 |
| 728149 | T-DM1 + Pertuzumab | 1 | 1 | 0 | 9.2248 | 8.3231 | 10.121 |
| 687288 | T-DM1 + Pertuzumab | 0 | 1 | 1 | 9.2631 | 9.9206 | 10.059 |
| 724293 | T-DM1 + Pertuzumab | 1 | 1 | 1 | 9.5155 | 8.1241 | 10.021 |

**Supplementary Table 2**

**RAB5A Allred score (IHC) of primary tumors from patients included from the KAMILLA study at Oslo University Hosptial. PFS (weeks). The data are blinded.**

| Patient# | PFS (weeks) | Proportion of positive stain score | Stain intensity score | Allred Score |
|---|---|---|---|---|
| 1 | 6.7 | 5 | 1 | 6 |
| 2 | 195.0 | 5 | 2 | 7 |
| 3 | 62.0 | 5 | 2 | 7 |
| 5 | 6.0 | 5 | 1 | 6 |
| 6 | 201.0 | 5 | 3 | 8 |
| 7 | 7.1 | 5 | 1 | 6 |
| 9 | 33.4 | 5 | 1 | 6 |
| 10 | 104.6 | 5 | 2 | 7 |
| 13 | 11.9 | 5 | 3 | 8 |
| 14 | 12.7 | 5 | 1 | 6 |
| 15 | 74.7 | 5 | 3 | 8 |
| 17 | 22.4 | 2 | 0 | 2 |
| 18 | 322.3 | 5 | 3 | 8 |
| 19 | 3.0 | 3 | 0 | 3 |
| 20 | 48.1 | 0 | 0 | 0 |
| 21 | 41.3 | 5 | 1 | 6 |
| 22 | 23.6 | 5 | 1 | 6 |
| 23 | 23.0 | 5 | 1 | 6 |
| 24 | 59.6 | 5 | 2 | 7 |
